# Clinical, dietary, and psychosocial effectiveness of interdisciplinary care in child and juvenile obesity

**DOI:** 10.1101/2020.06.09.20126953

**Authors:** Carlos Alberto Menezes, Rayzza Santos Vasconcelos

## Abstract

The aim of this study is to evaluate the effectiveness of interdisciplinary care in modifying anthropometric clinical parameters (BMI and waist circumference), psychosocial profile and eating behavior in a group of children and adolescents with obesity. This is a clinical study involving 200 children and adolescents with obesity who are participants of the “Serviço de Medicina Preventiva” program, and the “Centro de Especialidades Médicas de Crianças e Adolescentes” both in Aracaju, Sergipe, Brazil. At the end of the interdisciplinary intervention, there was a decrease in the number of hours devoted to electronic games and television and an increase in physical activity, which was associated with a reduction in reports of dissatisfaction and symptoms of depression. In addition, the sadness and prejudice related to obesity suffered at school and in the family environment were also attenuated, despite the percentage increase in the anxiety indicator. Regarding the anthropometric measurement, there was a reduction in body mass index, waist circumference, in caloric consumption. It is concluded that interdisciplinary programs are an effective strategy to reduce the negative repercussions of obesity in the daily lives of obese children and adolescents, as there were improvements in anthropometric parameters, in addition to psychosocial and dietary aspects.

**RESUMO:** O objetivo deste estudo é avaliar a eficácia do atendimento interdisciplinar na modificação de parâmetros clínicos antropométricos (IMC e circunferência da cintura), perfil psicossocial e comportamento alimentar em um grupo de crianças e adolescentes com obesidade. Trata-se de um estudo clínico envolvendo 200 crianças e adolescentes com obesidade participantes do programa “Serviço de Medicina Preventiva” e do “Centro de Especialidades Médicas de Crianças e Adolescentes”, ambos em Aracaju, Sergipe, Brasil. Ao final da intervenção interdisciplinar, houve uma diminuição no número de horas dedicadas a jogos eletrônicos e televisão e um aumento na atividade física, o que foi associado à redução nos relatos de insatisfação e sintomas de depressão. Além disso, a tristeza e o preconceito relacionados à obesidade sofrida na escola e no ambiente familiar também foram atenuados, apesar do aumento percentual no indicador de ansiedade. Em relação à medida antropométrica, houve redução do índice de massa corporal, circunferência da cintura e consumo calórico. Conclui-se que os programas interdisciplinares são uma estratégia eficaz para reduzir as repercussões negativas da obesidade no cotidiano de crianças e adolescentes obesos, pois houve melhorias nos parâmetros antropométricos, além de aspectos psicossociais e alimentares.

## INTRODUCTION

Obesity in the pediatric population is defined as body mass index above the 97th percentile of the body mass index/Center for Disease Control and Prevention (BMI/CDC) and is considered an important problem in public health with severe short- and long-term consequences for individuals, their families, and society.^(1)^ Children and adolescents who are overweight or obese have a higher risk to develop chronic diseases such as systemic arterial hypertension, type 2 diabetes mellitus, insulin resistance, dyslipidemia, metabolic syndrome, and certain types of cancer.^(2)^ In addition, obese children and adolescents also suffer from psychological repercussions such as body dissatisfaction, low self-esteem, depression, prejudice, and social stigmatization.^(3,4)^

The World Health Organization estimates that in European children, approximately 18-29% of them are overweight and 5-14% is obese. There are about 155 million overweight children and approximately 30 to 45 million obese children worldwide.^(5-7)^ Reference for Brazilian Studies show an overweight prevalence of: 10.1% in children and 20.5% in adolescents. In addition, a prevalence of obesity of 4.6% in children.^(8,9)^ The treatment of childhood obesity is a complex task that requires specific strategies aimed at this public, and it is imperative that families be included in the weight loss process.^(10)^ Interdisciplinary programs aimed at expanding the knowledge of childhood obesity have already been developed.^(11)^ However, studies on their effectiveness are still scarce. Treatments used in these programs are varied: single and group interventions, with or without medical and nursing supervision and familial, behavioral, and cognitive therapy, with or without prescription drugs. These variables are important goals to be adopted by the interdisciplinary team composed of endocrinologists, nutritionists, psychologists, physical educators, and social workers. Some programs are developed with opened or closed continued treatment, requiring incentives for individuals to participate regularly during the week, which restricts their practical applicability.^(12)^

Daniels^(13)^ directs the treatment of childhood obesity to behavioral changes and recommends that programs should establish permanent changes instead of short-term diets or exercise programs aimed at rapid weight loss. Management must include the family to make small, gradual behavioral and dietary changes on a daily basis.^(14)^ Further research is need to assess the varied consequences caused by this disease.

Among the repercussions of childhood obesity, attention is drawn to the need for studies on favorable clinical outcomes, as well as on the social, nutritional, and psychological behavior of obese pediatric patients. The aim of this study is to evaluate the effectiveness of interdisciplinary care in modifying anthropometric clinical parameters (BMI and waist circumference), psychosocial profile and eating behavior in a group of children and adolescents with obesity.

## MATERIALS AND METHODS

This is clinical study involving children and adolescents with obesity who are participants of the Preventive Medicine Service (SEMPRE), a private service; and the Center for Medical Specialties of Children and Adolescents (CEMCA), a public service, both in the city of Aracaju, in the state of Sergipe, Brazil. The study was approved by the Research Ethics Committee of the State University of Santa Cruz, under the presentation of Certificate for Ethics Assessment 04065412.600005526 of Platform Brazil.

The sample was composed of 200 children and adolescents attended at the CEMCA and SEMPRE institutions, comprising 100 males and 100 females, aged between 6 and 18 years. The inclusion criteria were children and adolescents who presented obesity and who were active participants in the reported public and private services programs. Exclusion criteria were psychiatric or hormonal disease verified in laboratorial exams and in the clinical routine of the medical service in SEMPRE or previous use of anti-obesity drugs. Participation of children was voluntary by applying a questionnaire and an informed consent form filled out and signed by the parents or guardians. Before starting the research, the purpose and essence of the study was explained to the participants. The participation of the subjects was by convenience and the results were submitted to power analysis. The study had a duration of 8 months (February / 2017 to October / 2017), without randomization and with loss of 10 subjects due to abandonment of the program.

The questionnaire was adapted and based on other instruments already established and validated such as the Pediatric Quality of Life Inventory and Measure health-related quality of life, and it is validated in other studies involving Brazilian population.^(15)^ The questionnaire was applied by doctors to program participants and their parent or guardian, during two 30-minute periods, at the beginning and again at the end of the programs, eight months after it had been initiated. The questionnaire approaches social and psychological aspects with the following items: lifestyle (hours per day on electronic games and television, psychological behavior, depression, anxiety, and sadness related to obesity), prejudice suffered at school, in the family environment and on the streets, along with aspects related with self confidence in response to the prejudice, satisfaction with family and school (physical activity), and body image. The study also covered whether these children and adolescents consider themselves ugly, whether they would like to be thin, and whether they feel ashamed of being obese and participating in a group with obese children. Other variables were included such as age, gender, ethnicity, education, family income, obesity in the family, and eating habits.

Initially, individuals were forwarded from Basic Health Units, linked to the Family Health Program with a previous diagnosis of childhood obesity established by the physician of the Basic Health Units. Next, they were submitted to a clinical evaluation by an endocrinologist and a nurse from CEMCA to confirm the diagnosis of obesity, followed by an evaluation by a nutritionist and a psychologist. From then on, the child’s parent or guardian was invited to participate in the program, whose main objective was to carry out activities related to learning how to eat healthy food, as well as activities related to the physical, psychological, and social aspects with children, adolescents, and their families. Meetings were held monthly at the program institutions and in public places. During the monthly meetings, children and adolescents, along with their parents or guardians, were clinically assessed by measuring weight (with digital anthropometer balance W200–100g Welmy–200kg Filizola, São Paulo, Brazil, in both public and private service, calibrated semi-annually by INMETRO/Lamas – with 4 sealers in total), height, BMI calculation, abdominal circumference, and blood pressure as shown by the algorithm in Figure 1.

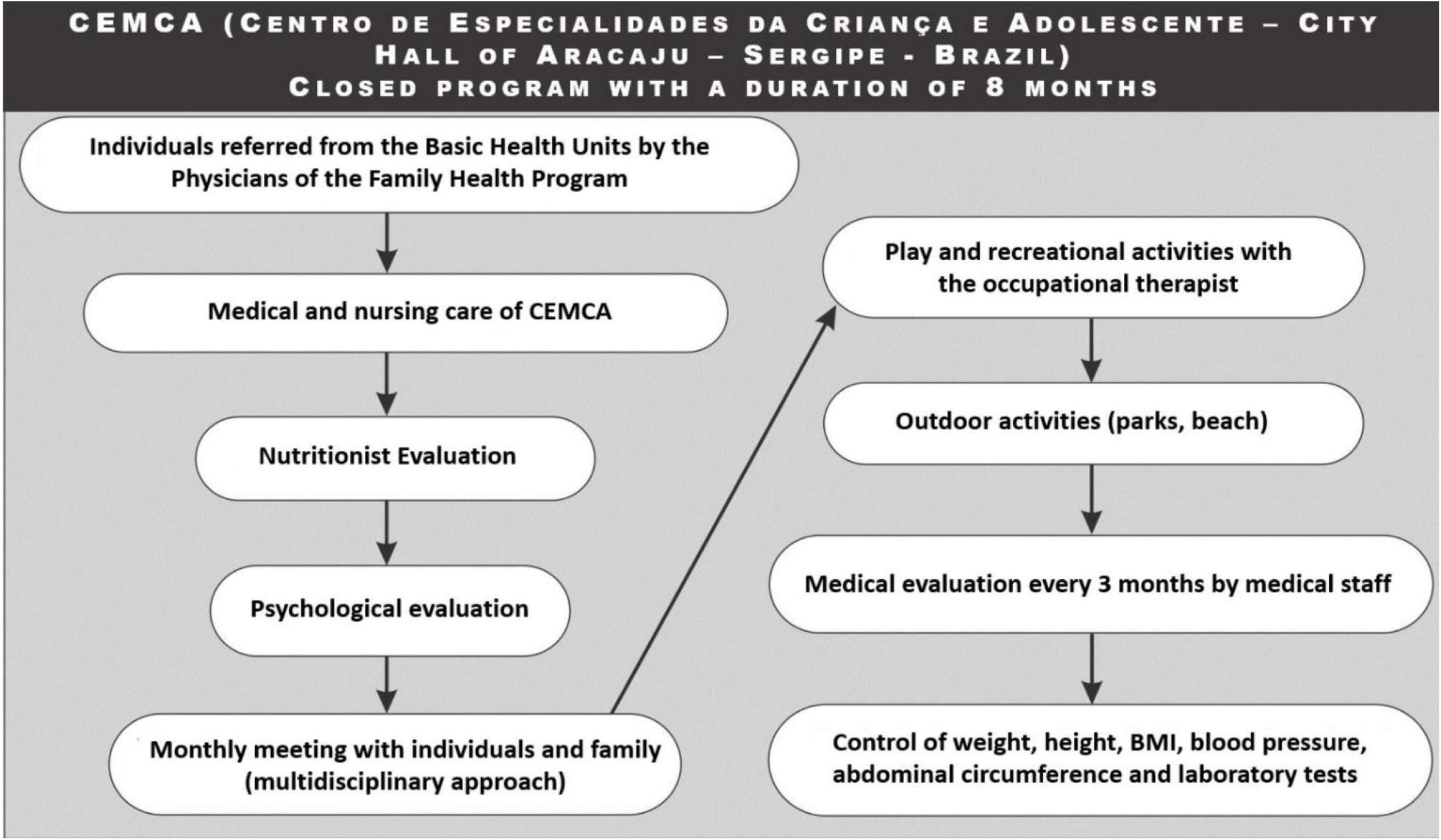

Conversely, individuals assisted at SEMPRE were referred by pediatric assistants from its offices. Next, they were submitted to a clinical evaluation by an endocrinologist and a nurse from SEMPRE to confirm the diagnosis of obesity, followed by an evaluation by a nutritionist and a psychologist. From then on, the child’s parent or guardian was invited to participate in the program, whose main objective was the same as CEMCA. Meetings were held weekly, and children, adolescents, and their guardian were clinically evaluated as shown by the algorithm in Figure 2.

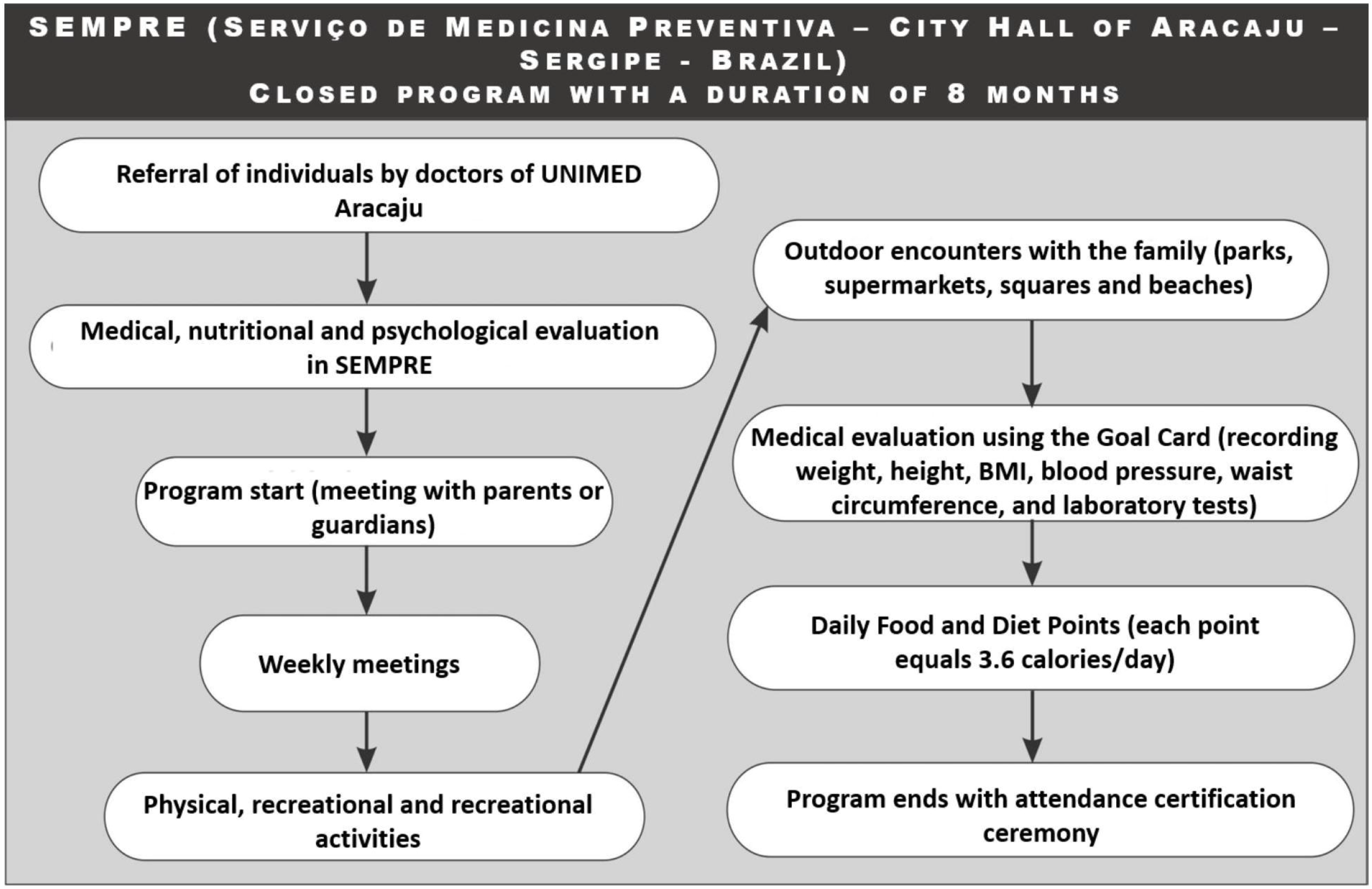

Therapy sessions were guided by a psychologist and conducted weekly with a group of children and a separate group of adolescents. Finally, meetings with nutritionists were individualized and the importance of food re-education was reinforced.

All individuals were clinically evaluated using anthropometric variables (weight and height) through the use of an anthropometric scale (Filizola^®^ Brazil). The BMI was used for characterizing obesity, followed by gender and age comparisons, according to the flowchart in the individual and group consultations as shown in Figures 1 and 2. According to the BMI table of CDC, a person is considered obese when the BMI is above the 97th percentile and overweight when the BMI is between 85-95 percentiles. The AC was also evaluated at the midpoint between the lower costal margin and the highest elevation of the iliac crest, and blood pressure was measured using the appropriate cuff for the pediatric group.

Individual behavior of the variables was evaluated at the beginning and at the end of eight months of the program in both public and private service groups. Quantitative variables were analyzed using the t-Student test. When the test assumptions were not fulfilled, the nonparametric Wilcoxon test was used. The main anthropometric and behavioral outcomes adjusted by baseline variables – physical activity, body mass index, abdominal circumference, caloric intake, and changes in dietary habits – were evaluated by analysis of covariance.

The Fisher’s exact test, McNemar test, and chi-squared test were used for qualitative variables and a difference was found between the beginning and the end of the program. To compare the effectiveness in both public and private services, a comparison was made between incidence of positive outcomes for both services by calculating the relative risk (RR), considering public service as the base.

## RESULTS AND DISCUSSION

The sample of 200 children who met the criteria for inclusion was distributed according to the genre. Among the male participants, the average age was 10.0±3.2 years and among the female participants, average age was 9.0±2.1years.Regarding skin color, 60% of all participants declared themselves as white in the private health service and 35% nonwhite in the public health service, and variable obesity of the parents was found in 83% of our sample in the public health service, and 62% in the private. Regarding the family income, the public service participants reported receiving two minimum wages and those of the private service, four. Finally, regarding eating behavior, 75% of the male gender was hyperphagic and 70% in the female gender was compulsive.

The questionnaire was self-applied and structured in the pattern of dichotomous questions and matrix response as shown in Table 1. The results of the participants’ social and psychological profiles in both services at the beginning and at the end of eight months of follow-up are presented in Table 1. Regarding the number of daily hours spent on electronic games and television, the individuals showed behavioral change, i.e., more children and adolescents spent less than 4 hours a day in both the public and the private services groups when evaluated at the end of the program (t=3.2; p<0.01). Reports of depression at the beginning of the program, with subsequent reduction at the end of the program were documented (t=2.6; p<0.01); conversely, there was an increase of anxiety levels at the end of the programs in both two groups (t=3.7; p<0.01).

**TABLE 1.** Results of social and psychological profile of obese children and adolescents at the beginning and after eight months at the end of the program. Aracaju, SE, 2017.

|  | SEMPRE<br>(n = 100) |  | CEMCA<br>(n = 100) |  | SEMPRE x CEMCA |
| --- | --- | --- | --- | --- | --- |
|  | Start (%) | End (%) | Start (%) | End (%) | p value |
| **Hours in electronic games and TV** |  |  |  |  |  |
| < 4 hours daily | 15 | 40 | 10 | 40 | <0.01† |
| 4 to 6 hours daily | 50 | 50 | 40 | 30 | <0.01† |
| > 6 hours daily | 35 | 10 | 50 | 30 | <0.01† |
| **Psychological behavior** |  |  |  |  |  |
| Depression | 25 | 15 | 20 | 10 | <0.01* |
| Anxiety | 25 | 48 | 14 | 50 | <0.01* |
| Sadness related to obesity | 50 | 37 | 66 | 40 | <0.001* |
| **Prejudice** |  |  |  |  |  |
| School | 50 | 20 | 45 | 15 | <0.001* |
| Family | 22 | 15 | 18 | 11 | <0.001* |
| Street | 20 | 10 | 31 | 21 | <0.01* |
| Self confidence | 8 | 55 | 6 | 53 | <0.001* |
| **Satisfaction with family** |  |  |  |  |  |
| Very Satisfied | 31 | 50 | 25 | 40 | <0.01* |
| Satisfied | 24 | 30 | 31 | 35 | <0.05* |
| Dissatisfied | 45 | 20 | 44 | 15 | <0.01* |
| **Satisfaction with School (Physical activity)** |  |  |  |  |  |
| Very Satisfied | 25 | 35 | 33 | 40 | <0.01* |
| Satisfied | 35 | 45 | 30 | 32 | <0.05* |
| Dissatisfied | 40 | 20 | 37 | 28 | <0.01* |
| **Body Image** |  |  |  |  |  |
| Feel ugly | 1 | 1 | 1 | 1 | 0.213 |
| Would like to be thin | 30 | 12 | 20 | 15 | <0.001* |
| Do not like to look in the mirror | 11 | 5 | 9 | 8 | <0.05* |
| Ashamed of being obese | 40 | 21 | 35 | 18 | <0.001* |
| Experience in group | 18 | 61 | 25 | 58 | <0.001* |

Source: Research data.
Subtitle: CEMCA (Center for Medical Specialties of Children and Adolescents)
SEMPRE (Preventive Medicine Service)
p-value (p- value at the beginning and at the end of programs)
\*(Fisher exacttest)
†(McNemartest)

Prejudice suffered by participants was more related to family and school, but a reduction of this variable was observed, in addition to better self-confidence at the end of the program in both service (t=4.3; p<0.001). Regarding the profile of satisfaction related to the family and school (physical activity), the participants showed dissatisfaction both in the public the private service (t=3.7; p<0.01) at the beginning of the program, but a subsequent feeling of satisfaction at the end (t=2.6; p<0.01).

This initial dissatisfaction reflects the prejudice suffered in these environments for both health service. Regarding body image, most of the participants wanted to be thin at the end of the program with (t=4.3; p<0.001) and did not like their image on the mirror. Despite these feelings experienced at the beginning of the programs, they had a better body image evaluation at the end of the programs (t=2.44; p<0.05). Therefore, children and adolescents had self-esteem because they were obese and showed improvement at the end of the study (t=4.4; p<0.001). Finally, there was an increase in satisfaction in group experiences comparing beginning and the end of both the public and private programs (t=4.3; p<0.001) as shown in Table 1.

A comparison between favorable outcome variables depicting the effect of the programs in both public and private health services at the beginning and end of eight months is shown in Table 2.

**TABLE 2.** Comparison of the favorable outcome variables.

| Variables analyzed | RR | 95% CI | p value |
| --- | --- | --- | --- |
| ↑ Sports practice | 0.8 | (0.552-1.919) | 0.03* |
| ↓ BMI (kg/m<sup>2</sup>) | 0.22 | (0.650-2.020) | 0.021* |
| ↓ Abdominal circumference (AC) | 0.40 | (0.430-3.050) | 0.03† |
| ↓ Caloric intake | 0.53 | (0.312-1.380) | 0.023* |
| ↑ Changing eating habits. | 0.64 | (0.320-1.230) | 0.212* |

Source: Research data.
Subtitle: BMI (body mass index)
RR (relative risk)
p-value (p-value at the beginning and at the end of programs)
CI (confidence interval)
Caloric intakes (kcal needed for sex and age - intake)/intake
\* “qui- quadrado test”
† (Fisher exact test)
↑ (Elevation)
↓ (reduction)

The RR shown is related to the two groups based on the public service when compared to the beginning and the end of the programs. In relation to the practice of sports and there was a change in healthy eating habits, there was an increase in RR, on the other hand the BMI, AC and caloric intake had a reduction in RR. The results show no difference between the two groups. (Table 2)

The amount of food per monthly consumption eby portions, units, cups, and the frequency analysis is shown in Table 3, depicting the eating behavior of the individuals of both groups and revealing positive changes in diet at the end of the assistance programs carried out by the interdisciplinary team. (Table 3)

**TABLE 3.** Food ingested at the beginning and end of the eight months.

|  | Start | End | p value |
| --- | --- | --- | --- |
| ↑ Fruits and vegetables (units) | 30 | 60 | 0.002* |
| ↓ Carbohydrates (serving) | 140 | 120 | 0.341 |
| ↓ Proteins (serving) | 45 | 40 | 0.328 |
| ↓ Fats (serving) | 100 | 70 | 0.002* |
| ↑ Water and juices (cups) | 138 | 199 | 0.003* |
| ↓ Soda (cups) | 30 | 10 | 0.002* |
| ↓ Chips and fast food (units) | 22 | 15 | 0.001* |
| ↓ Sweets and ice cream(units) | 20 | 10 | 0.002* |
| ↑ Milk and milk derivatives (cups) | 27 | 37 | 0.003* |

Source: Research data.
Subtitle: p-value (p- value at the beginning and at the end of programs)
\* t Student test
↑ (Elevation)
↓ (Reduction)
\* Wilcoxon Test

Epidemiological studies have shown the role that obesity plays in the development of the psychological and behavioral profile of children and adolescents, ^(1,3,4)^ demonstrating that weight gain can have negative consequences in relation to the self-image and emotional relationships of these individuals. In this study, we identified dissatisfaction in family and social relationships, prejudice, body self-image fragmentation and reporting of physical activity practices in obese children and adolescents at the beginning and at the end of eight months.

Obesity is related to reports of depression, anxiety, loneliness and behavioral changes. These can interfere in the child’s life, impairing school performance and family and social relationships. ^(5,6,10,11)^

In this context, and corroborating with other studies, it was verified that children and adolescents presented symptoms of depression and most reported feeling sad because they were obese at the beginning of the program, which were reduced at the end, while there was an increase in anxiety in the end of the period in both services. Puder et al. found prevalence of significant depression in children, from 0.4 to 3.0%, and from 3.3 to 12.4% in adolescents with obesity, when compared to normal controls.^(14)^ In this study, this variable was not compared in non-obese children and adolescents. However, there is no consensus in the literature on the relationship between depression and obesity.^(13)^

Discrimination among children can have serious psychological implications. Pearce et al., in a study of 416 high school students, found high levels of discrimination among children and adolescents with obesity.^(16)^ Children who suffered prejudice and discrimination by appearance had high rates of body dissatisfaction, depression and low self-esteem.^(16-18)^

In this study, it was revealed that the majority of the children and adolescents suffered some type of prejudice, being the school and family environment the most correlated places, regardless of the medical service in which they participated.

Several studies have shown dissatisfaction in the relationship between obese children and their families, since they are forced to practice physical activities and are prevented from eating high calorie foods, eventually developing a conflictive relationship with their parents.^(18.19)^These studies showed that participants were dissatisfied with family life because of the high degree of prejudice they had, and experienced a reduction in their dissatisfaction after the completion of interdisciplinary programs.^(14)^

The dissatisfaction among the individuals of the public and private school institutions with the practice of physical activity at the beginning of the programs was also demonstrated; at the end of these programs, there was an increased need to practice sports, confirming findings from studies 13 that demonstrate that overweight children do less physical activity because they feel discriminated during these activities by their appearance. Therefore, it is necessary to develop public policies to encourage physical activity and healthy eating.^(20-25)^

In relation to eating habits, male children and adolescents are hyperphagic and women are compulsive, reflecting the high degree of prejudice they suffer.^(1,8)^ This fact reflects a greater consumption of snack foods and fast food, carbohydrates, fats and soft drinks, but at the end of the programs in the public and private services, there was a reduction in the consumption of these hypercaloric foods and an increase in the consumption of healthy foods, positively affecting the reduction of the BMI and AC, as observed in our study.^(25)^ Another important report was related to the excess of spent hours in electronic games and television up to 6 hours a day, as confirmed by Thompson et al.^(20)^ In addition, television can develop conflicting thoughts because the same instrument that explores healthy eating habits is able to stimulate the consumption of fast food.^(24)^

Therefore, it is necessary to introduce healthy eating habits as soon as possible, helping obese children and adolescents to avoid metabolic pathologies in adult life.^(21.22)^ Finally, it is necessary to organize interdisciplinary care programs to control pediatric obesity, due to the multiple interfaces that this disease presents. It is possible to compare individualized outpatient care with group education programs addressing changes in eating habits and physical activity and to conclude that both obesity management strategies favored changes in eating habits and physical activity level. Group medical care in a nutritional and health education program has been the same or more effective than individualized follow-up and should be seen as an alternative for the treatment of childhood obesity.^(12)^

In our study, we did not compare individualized and interdisciplinary care, but we compared the interdisciplinary care between two public and private medical services with different social and economic realities, but with the same problem: obesity. In our comparison, the following observations were highlighted: high incidence of maternal and paternal obesity, with higher levels in the public service (83%) in relation to private service (62%); an average family income of 2 minimum wages in the public service and 4 minimum wages in the private service. The predominance of compulsive psychological behavior of girls and hyperphagia in boys was evidenced in both services.

In addition, the improvement of the social, behavioral and psychological patterns of these individuals occurred in both services, with no differences between these two groups at the end of the study.

In this context, we recommend treatment of pediatric obesity by interdisciplinary teams, due to the beneficial effects on indicators of obesity and its psychosocial consequences.

In conclusion, obesity has a negative influence on the psychosocial development of children and adolescents, especially with regard to self-perception of the body, feelings of depression, sadness, anxiety and low self-esteem, all of which reflect a compromised quality of life. This study demonstrated that the interdisciplinary care of obese children and adolescents improves clinical parameters such as BMI and AC, as well as the psychosocial profile and eating habits when comparing individuals at the beginning and at the end of programs offered by public and private health services.

## Data Availability

We inform that all the data of the manuscript are available at the Preventive Medicine Service and at the Center for Medical Specialties of Children and Adolescents

